# Evaluating the Effect of Inequalities in Oral Anti-coagulant Prescribing on Outcomes in People with Atrial Fibrillation

**DOI:** 10.1101/2023.08.28.23294755

**Authors:** R.J Mulholland, F. Manca, G. Ciminata, T.J Quinn, R. Trotter, K.G. Pollock, S. Lister, C. Geue

## Abstract

**Background:** Whilst anti-coagulation is typically recommended for thromboprophylaxis in atrial fibrillation (AF), it is often never prescribed, or prematurely discontinued, due to concerns regarding bleeding risk. The aim of this study was to assess both stroke/systemic embolism (SSE) and bleeding risk, comparing people with AF who continue anticoagulation with those who stop transiently, stop permanently or never start.

**Methods:** This retrospective cohort study utilised linked Scottish administrative healthcare data to identify adults diagnosed with AF between January 2010 and April 2016, with a CHA_2_DS_2_- VASC score of ≥2. They were sub-categorised into cohorts based on anti-coagulant exposure: never started, continuous, discontinuous, and cessation. Inverse probability of treatment weighting-adjusted Cox regression and competing-risks regression were utilised to compare the risks of SSE and major bleeding between cohorts during a five year follow-up period. Sub-group analyses evaluating risk of SSE, bleeding and mortality, were undertaken for people commenced on anti-coagulation that experienced a major bleeding event

**Results:** Of an overall cohort of 47,427 people, 26,277 (55.41%) were never anti-coagulated, 7,934 (16.72%) received continuous anti-coagulation, 9,107 (19.2%) temporarily discontinued and 4,109 (8.66%) permanently discontinued. Initiation and continuation of anti-coagulation was less likely in people with a lower socio-economic status, elevated frailty score, or aged ≥75. SSE risk was significantly greater in those with discontinuous anti-coagulation, compared to continuous (SHR: 2.65; 2.39-2.94). In the context of a major bleeding event, there was no significant difference in bleeding risk between the cessation cohort compared to those that continued anti-coagulation (SHR 0.94; 0.42-2.14).

**Conclusion:** Our data suggest significant inequalities in anti-coagulation prescribing for people with AF, with substantial opportunity to improve initiation and continuation. Anti-coagulation decision-making must be patient-centered and recognise that discontinuation or cessation is associated with a substantial risk of thromboembolic events not offset by a reduction in bleeding.

**What is Known?:**

- Despite a high thromboembolic risk, anti-coagulation in people with atrial fibrillation is frequently not initiated, or prematurely discontinued

**What is New?:**

- Our data suggest considerable inequalities in anti-coagulation prescribing in people with atrial fibrillation; people with a lower socio-economic status, elevated frailty score, or aged ≥75 were less likely to initiate or continuation anti-coagulation
- Whilst non-initiation and cessation of anti-coagulation are associated with elevated thromboembolic risk, this risk is particularly high in people with atrial fibrillation that transiently discontinue anti-coagulation
- In the context of a major bleeding event, permanent discontinuation of anti-coagulation in people with atrial fibrillation is not associated with a significantly reduced risk of recurrent bleeding compared to those that are continuously anti-coagulated.

## Introduction

### Background/Rationale

Despite the high thromboembolic risk in the AF population, stroke prophylaxis with anti-coagulants is frequently under-utilised or prematurely discontinued, generally due to the monitoring requirements of warfarin or a perceived high risk of bleeding.(1) Indeed, the likelihood of older adults and females being anti-coagulated is paradoxically lower, despite their higher stroke risk.(2) A prior observational study of people with non-valvular AF (NVAF) prescribed warfarin in a national dataset combining Medicare and insurance claims data, reported that the risk of ischaemic stroke is approximately doubled in those that discontinued warfarin compared to those with continuous prescriptions.(3) A study of 1361 individuals with NVAF prescribed an alternative vitamin K antagonist, acenocoumarol, at an anti-coagulation centre in Spain suggested that cessation of anti-coagulation is associated with increased stroke, adverse cardiovascular events and all-cause mortality.(4) Whilst the acute treatment of those with bleeding associated with anti-coagulation often requires immediate discontinuation of anti-coagulation, the ongoing management is complex; clinical decision-making must consider the competing risks of a thromboembolic event if anti-coagulation is withheld, and a recurrent bleed if it is recommenced, and clinical consensus is currently lacking as to the optimal approach. (5) Whilst there is an increasing body of literature around patterns of adherence to anti-coagulation, few studies have evaluated the clinical outcomes associated with discontinuation of anti-coagulation in individuals with AF, particularly in the context of a major bleeding event.

All individuals in Scotland are assigned a unique identification number, the Community Health index (CHI) number, creating a record of engagements with health and social care facilities through the lifetime. (6, 7)Linkage of national databases by CHI number affords the opportunity to analyse rich Scotland-wide individual patient data; this real-world data may be leveraged to explore research questions, such as the impact on clinical outcomes of discontinuing oral anti-coagulation in people with AF for which a randomised-controlled trial may be infeasible.(8–11)

### Objectives

The primary objective of this study was to compare the risks of stroke/systemic embolism (SSE) in adults with AF with discontinuous exposure to anti-coagulation, versus those never started on anti-coagulation, and those with continuous anti-coagulation, respectively. The secondary objective was to compare the risk of SSE and bleeding in those that received continuous OAC therapy with those that discontinued anti-coagulation in the context of a major bleeding event.

## Methods

### Study Design

This was a retrospective observational cohort study of adults hospitalised with an incident AF event in Scotland.

### Data Sources and Cohort

Methods are reported in accordance with the REporting of studies Conducted using Observational Routinely-collected Data (RECORD) guidelines (Supplementary Materials).(12) Public Health Scotland (PHS) provided access to fully anonymised data in support of a broader study which utilised routinely collected healthcare data to assess the comparative effectiveness of anticoagulation for stroke prevention in people diagnosed with AF.

All adults aged 18 or older with a diagnosis of AF between 1^st^ January 2010 and April 30^th^ 2016 with a CHA_2_DS_2_-VASC score of two or greater were identified from Scottish Morbidity Records (SMR) 01, which records inpatient and day case discharges for all specialities excluding psychiatry and obstetrics. We triangulated this with data from the Scottish Stroke Care Audit (SSCA), which collected information from all Scottish hospitals managing stroke care and includes data on demographics, and AF diagnosis and management. Agreement between the datasets is outlined in Supplementary Material 3. The AF cohort in SMR01 was delineated by ICD-10 coding in any diagnostic position(I48). Patient-level data linkage was undertaken with National Records of Scotland (NRS) (mortality data), Prescribing Information System(PIS) (prescribing data) and SMR00 (outpatient appointments) (Figure 1). Stroke and bleeding events were measured using International Classification of Diseases (ICD)-10 and Office of Population Censuses and Surveys (OPCS)-4 codes (Supplementary Materials: Table S2).(13) Data were available from 1^st^ January 2005, allowing for a five-year lookback period to identify those whose primary AF diagnosis preceded 1^st^ January 2010. Data was available until 31^st^ May 2021, and individuals were followed-up until the event of interest and were censored at the end of the follow-up period, five years after the index date of AF diagnosis.

**Figure 1.**
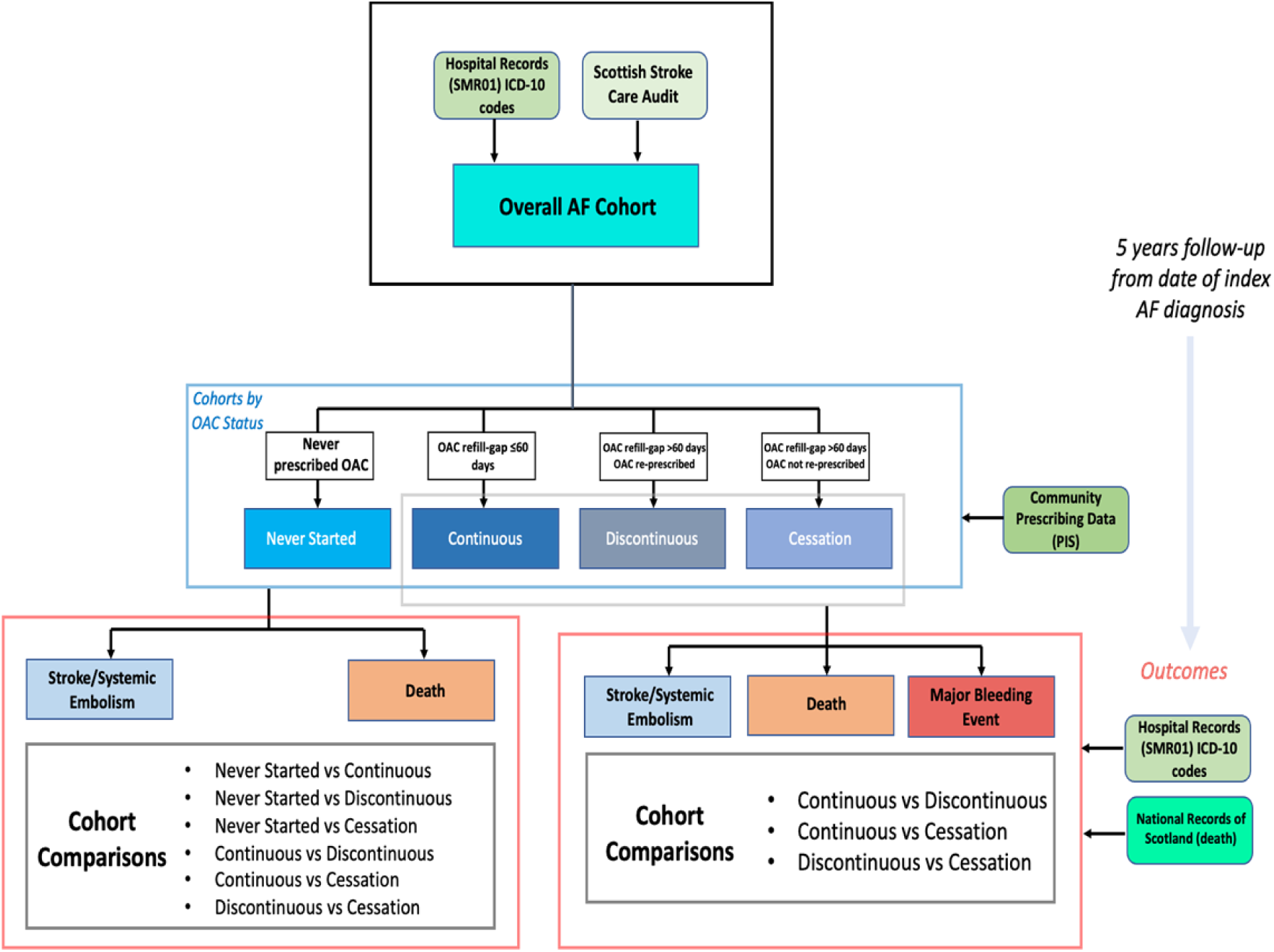
Outline of identification of AF population and formation of cohorts utilised in analyses, including sub-group analyses.

Our AF population was divided into four cohorts according to exposure to oral anti-coagulation: ‘never started’, ‘continuous OAC therapy’, ‘discontinuous OAC therapy,’ and ‘cessation of OAC therapy,’ (Table 1). Anti-coagulation is here defined as having been prescribed either warfarin or a DOAC (apixaban, rivaroxaban, dabigatran or edoxaban) for the first time following an AF event. Anti-coagulation prescriptions with dosages less than a minimum therapeutic threshold for prevention of thromboembolic events defined by the British National Formulary (BNF) were excluded. A lookback period of four-years prior to respective index AF events was implemented to ensure only individuals who were treatment naïve prior to commencing anti-coagulation were included in the analysis.

**Table 1.** Cohort Definitions.

|  |  |
| --- | --- |
| Never Started | Individuals with an incident AF event during the study that were not prescribed an oral anti-coagulant |
| Continuous OAC Therapy | Individuals with an incident AF event during the study that were prescribed an oral anti-coagulant with no refill gaps exceeding the defined threshold for discontinuation |
| Discontinuous OAC Therapy | Individuals with an incident AF event during the study that were prescribed an oral anti-coagulant, with a refill gap exceeding the defined threshold for discontinuation. |
| Cessation of OAC Therapy | Individuals with an incident AF event during the study that were prescribed an oral anti-coagulant, with a refill gap exceeding the defined threshold for discontinuation, that receive no further prescriptions of an oral anti-coagulant during the observation period. |

Discontinuation of anti-coagulation was established according to the refill-gap method; those with a temporal gap between consecutive prescriptions of greater than sixty days, not filled by the penultimate prescription, were defined as having discontinued.(14) Prescriptions of oral anti-coagulants were determined from the PIS; this dataset comprises records for all prescribed drugs dispensed by community pharmacies, in addition to dispending physicians and suppliers of medical devices.(15) To ensure that the indication for the anti-coagulant prescription was exclusively AF, Individuals with valvular heart disease, a mechanical cardiac valve, or venous thromboembolism, were excluded from analyses (Table S3).

Sub-group analyses evaluating risk of SSE, bleeding and mortality, were undertaken for people commenced on anti-coagulation that experienced a major bleeding event.

Data cleaning and pre-processing was undertaken according to established methodologies for routine healthcare data.(16) Since less than 5% of records had missing data, and these appeared to be missing completely at random, imputation of the missing values was not completed; instead, a complete case analysis was undertaken, with those records excluded. Variables with missing data are detailed in Appendix 3 (Table S4).

The propensity score-based Inverse Probability of Treatment Weighting (IPTW) method was utilised to address potential confounding by indication, arising due to lack of randomisation.(17) IPTW was utilised rather than propensity score matching to avoid the exclusion of eligible subjects. Propensity score (PS) estimation was undertaken for the respective AF subgroups to estimate the probability of anti-coagulant exposure status conditional upon the observed baseline characteristics of the AF population.(18) Up to five PS models (never started versus discontinuous OAC therapy, never started versus cessation of OAC therapy, discontinuous OAC therapy versus cessation of OAC therapy, continuous OAC therapy versus discontinuous OAC therapy, continuous OAC therapy versus cessation of OAC therapy) were thus generated for each outcome of interest. Logit models were used to estimate propensity scores and incorporated baseline characteristics which were either of prognostic relevance to the clinical outcomes, or potentially predictive of anti-coagulation status. The models accounted for age (years), sex, and geographical location using an 8-fold urban rurality classification, which was divided into three strata: urban (1–2), small and large towns (3–6), and rural (7–8). An indicator of socio-economic status, the Scottish Index of Multiple Deprivation (SIMD) was also included; this is a ranked scale of multiple deprivation for geographical locations, and was divided into quintiles, such that one represents the most deprived, and five denotes the least deprived areas, respectively. PS also accounted for CHA_2_DS_2_-VASc at the time of the index AF event. Furthermore, PS for the ischaemic stroke/systemic embolism outcomes also accounted for the following confounding variables: prior stroke/TIA, comorbidity (measured using the Charlson Co-morbidity Index), and an electronic frailty score. (19) PS for the outcomes for major bleeding also accounted for anti-platelet prescriptions in the two years preceding the index AF event and time (days).

To implement the IPTW methodology, a weight indicative of the probability of either no exposure to anti-coagulation or continuous OAC therapy, and identical to the reciprocal of the aforementioned PS, was applied to individuals in those respective cohorts.(18) Similarly, a weight equivalent to the reciprocal of one minus the PS was allocated to the discontinuous OAC therapy cohort. The risks of two significant clinical events associated with AF, stroke (both ischaemic and haemorrhagic), major bleeding between discontinuation of anti-coagulation in individuals diagnosed with AF, was compared for those never prescribed anti-coagulation, and for those with continuous exposure, using Cox proportional hazards regression (Figure 2). Doubly robust estimation was implemented by inclusion of the aforementioned propensity scores within the respective Cox proportional hazards regression models. To mitigate bias due to residual differences in the baseline covariates, and redress possible covariate imbalance, the variables included to estimate propensity scores were incorporated into the adjusted models. Competing risk regression, using Fine & Gray proportional sub-hazards models, was implemented to establish the first clinical event (SSE or death, and major bleed or death). Outcomes were evaluated using an intention-to-treat analysis.

**Figure 2.**
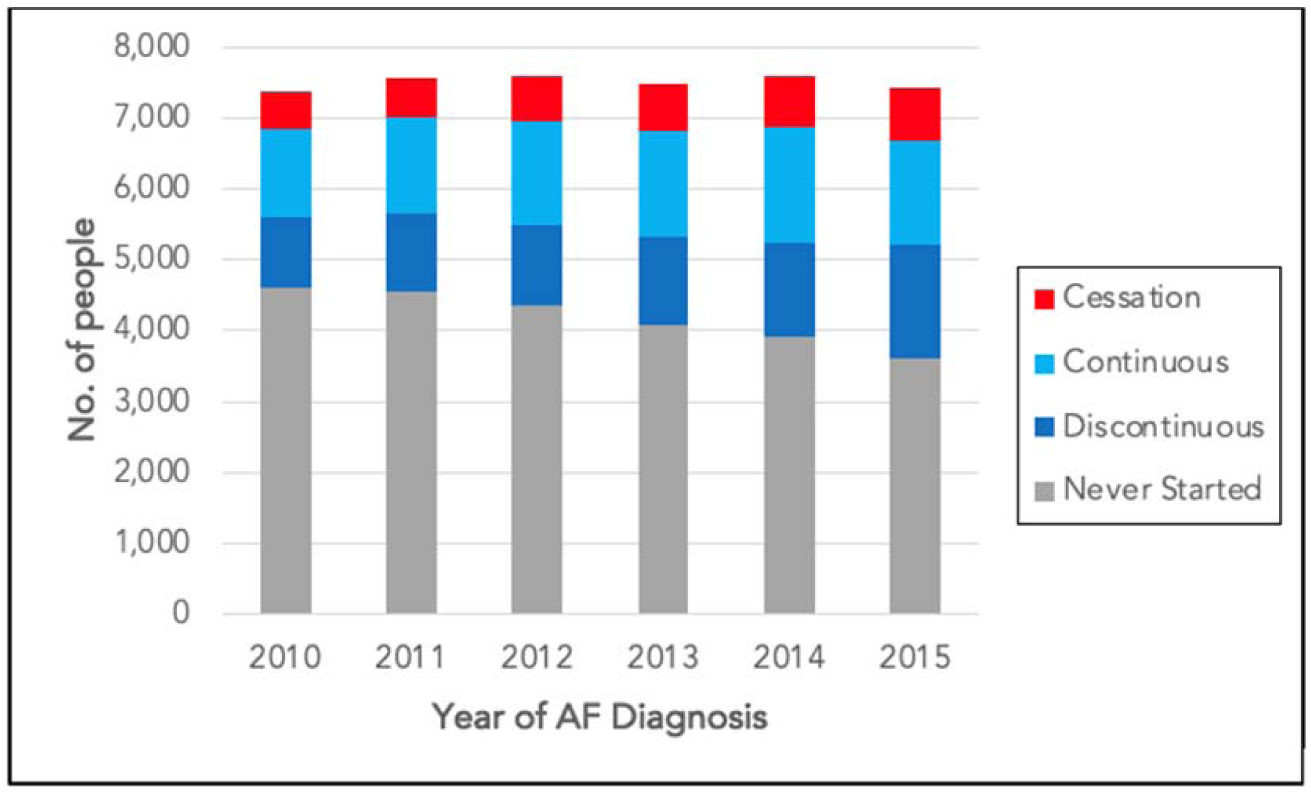
OAC exposure by year of AF diagnosis

Statistical analyses were completed using STATA version 16. Descriptive statistics for categorical variables are presented as the frequency and percentage in which they occurred. Regression analyses are presented as hazard ratios with 95% confidence intervals.

### Ethics

The study did not require review and approval by an NHS Research Ethics Committee since it utilised de-identified routinely collected healthcare data. This study was conducted in accordance with International Society for Pharmacoepidemiology (ISPE) Guidelines for Good Pharmacoepidemiology Practices (GPP) and applicable regulatory requirements.

### Data Availability

The data is confidential, and analyses were subject to disclosure control by PHS. Access to the data may be requested by application to PHS via the Public Benefit and Privacy Panel.

## Results

### Characteristics of the AF Cohort

Of an overall cohort of 47,427 people with an incident diagnosis of AF between 1^st^ January 2009 and 30^th^ April 2016 and a CHA_2_DS_2_-VASC score of ≥2, those never prescribed anti-coagulations tended to be older, comprising a considerably greater proportion of people aged older than 85 (Table 2). Sex, urban rurality score, CHA_2_DS_2_-VASc scores, and HAS-BLED scores were comparable between the cohorts. The cohort that never initiated anti-coagulant therapy had the greatest proportion of individuals in the most deprived SIMD quintile, whilst the continuous OAC therapy population had the greatest proportion of people categorised within the most affluent quintile. Furthermore, those aged ≥75, or with an elevated frailty score, were less likely to initiate anti-coagulation, and if prescribed, were more likely to discontinue.

**Table 2.** Descriptive statistics by anti-coagulation status.

|  |  | **Never**<br>**Started**<br>**(N=26277)** | **Continuous**<br>**(N=7934)** | **Discontinuous**<br>**(N=9107)** | **Cessation**<br>**(N=4109)** |
| --- | --- | --- | --- | --- | --- |
| **Age** | ≤55 | 429 | 188 | 208 | 65 |
|  | % | 1.63 | 2.37 | 2.28 | 1.58 |
|  | 56-75 | 6,311 | 3,620 | 4,216 | 1,213 |
|  | % | 24.02 | 45.63 | 46.29 | 29.52 |
|  | 76-85 | 10,634 | 3,242 | 3,862 | 1,960 |
|  | % | 40.47 | 40.86 | 42.41 | 47.70 |
|  | >85 | 8,903 | 884 | 821 | 871 |
|  | % | 33.88 | 11.14 | 9.02 | 21.20 |
| **Sex** | Female | 15,182 | 4,496 | 5,141 | 2,202 |
|  | % | 57.78 | 56.67 | 56.45 | 53.59 |
|  | Male | 11,095 | 3,438 | 3,966 | 1,907 |
|  | % | 42.22 | 43.33 | 43.55 | 46.41 |
| **CHA<sub>2</sub>DS<sub>2</sub>-**<br>**VASC Score** | 2 | 6,404 | 2,292 | 2,970 | 994 |
|  | % | 24.37 | 28.89 | 32.61 | 24.19 |
|  | 3 | 9,041 | 2,378 | 2,848 | 1,286 |
|  | % | 34.41 | 29.97 | 31.27 | 31.30 |
|  | 4 | 6,066 | 1,684 | 1,822 | 924 |
|  | % | 23.08 | 21.23 | 20.01 | 22.49 |
|  | 5 | 3,135 | 1,035 | 985 | 601 |
|  | % | 11.93 | 13.05 | 10.82 | 14.63 |
|  | 6 | 1,255 | 428 | 373 | 224 |
|  | % | 4.78 | 5.39 | 4.10 | 5.45 |
|  | 7 | 322 | 107 | 100 | 68 |
|  | % | 1.23 | 1.35 | 1.10 | 1.65 |
|  | 8 | 47 | 10 | 9 | 12 |
|  | % | 0.18 | 0.13 | 0.10 | 0.29 |
|  | 9 | 7 | 0 | 0 | 0 |
|  | % | 0.03 | 0.00 | 0.00 | 0.00 |
| **HAS-BLED**<br>**Score** | 0 | 121 | 53 | 69 | 20 |
|  | % | 0.46 | 0.67 | 0.76 | 0.49 |
|  | 1 | 4697 | 1628 | 1756 | 789 |
|  | % | 17.87 | 20.52 | 19.28 | 19.2 |
|  | 2 | 10236 | 3090 | 3736 | 1529 |
|  | % | 38.95 | 38.95 | 41.02 | 37.21 |
|  | 3 | 7660 | 2300 | 2594 | 1222 |
|  | % | 29.15 | 28.99 | 28.48 | 29.74 |
|  | 4 | 2899 | 744 | 827 | 476 |
|  | % | 11.03 | 9.38 | 9.08 | 11.58 |
|  | ≥5 | 664 | 119 | 125 | 73 |
|  | % | 2.53 | 1.5 | 1.37 | 1.78 |
| **Grouped**<br>**Charlson**<br>**Index** | 0 | 10,031 | 4,223 | 4,797 | 1,870 |
|  | % | 38.17 | 53.23 | 52.67 | 45.51 |
|  | 1 | 5,407 | 1,660 | 1,985 | 865 |
|  | % | 20.58 | 20.92 | 21.80 | 21.05 |
|  | 2 | 10,839 | 2,051 | 2,325 | 1,374 |
|  | % | 41.25 | 25.85 | 25.53 | 33.44 |
| **Urban**<br>**Rurality** | 1-2 (City) | 18,620 | 5,374 | 6,204 | 2,797 |
|  | % | 70.86 | 67.73 | 68.12 | 68.07 |
|  | 3-6 (Towns) | 3,337 | 1,057 | 1,205 | 550 |
|  | % | 12.70 | 13.32 | 13.23 | 13.39 |
|  | 7-8 (Rural) | 4,320 | 1,503 | 1,698 | 762 |
|  | % | 16.44 | 18.94 | 18.64 | 18.54 |
| **SIMD** | 1 | 6,153 | 1,688 | 1,933 | 857 |
|  | % | 23.42 | 21.28 | 21.23 | 20.86 |
|  | 2 | 6,328 | 1,796 | 2,038 | 996 |
|  | % | 24.08 | 22.64 | 22.38 | 24.24 |
|  | 3 | 5,349 | 1,642 | 1,946 | 911 |
|  | % | 20.36 | 20.70 | 21.37 | 22.17 |
|  | 4 | 4,552 | 1,476 | 1,689 | 708 |
|  | % | 17.32 | 18.60 | 18.55 | 17.23 |
|  | 5 | 3,895 | 1,332 | 1,501 | 637 |
|  | % | 14.82 | 16.79 | 16.48 | 15.50 |
| **Prior Stroke** | % | 1,615 | 367 | 346 | 171 |
|  |  | 6.15 | 4.63 | 3.80 | 4.16 |
| **Previous**<br>**anti-platelet** | % | 15523 | 4146 | 5134 | 2318 |
|  |  | 59.07 | 52.26 | 56.37 | 56.41 |
| **Frailty Score** | Low risk <5 | 7,114 | 4,204 | 4,174 | 1,045 |
|  |  | 27.07 | 52.99 | 45.83 | 25.43 |
|  | Moderate<br>risk 5-15 | 10,081 | 2,679 | 3,172 | 1,543 |
|  |  | 38.36 | 33.77 | 34.83 | 37.55 |
|  | High risk >15 | 9,082 | 1,051 | 1,761 | 1,521 |
|  |  | 34.56 | 13.25 | 19.34 | 37.02 |

The rate of discontinuation appears to be greatest at the extremes of the CHA_2_DS_2_-VASC score, with rates of 63.4% and 63.5% for scores of 2 and 3, and 63.6% and 67.7% for scores of 7 and 8 respectively. Initiation and continuity of anti-coagulation improved according to year of AF diagnosis (Figure 2). DOACs replaced warfarin as the predominant OAC prescribed over the duration of the study (Figure 3) although OAC discontinuity was comparable between OAC types (Figure S1).

**Figure 3.**
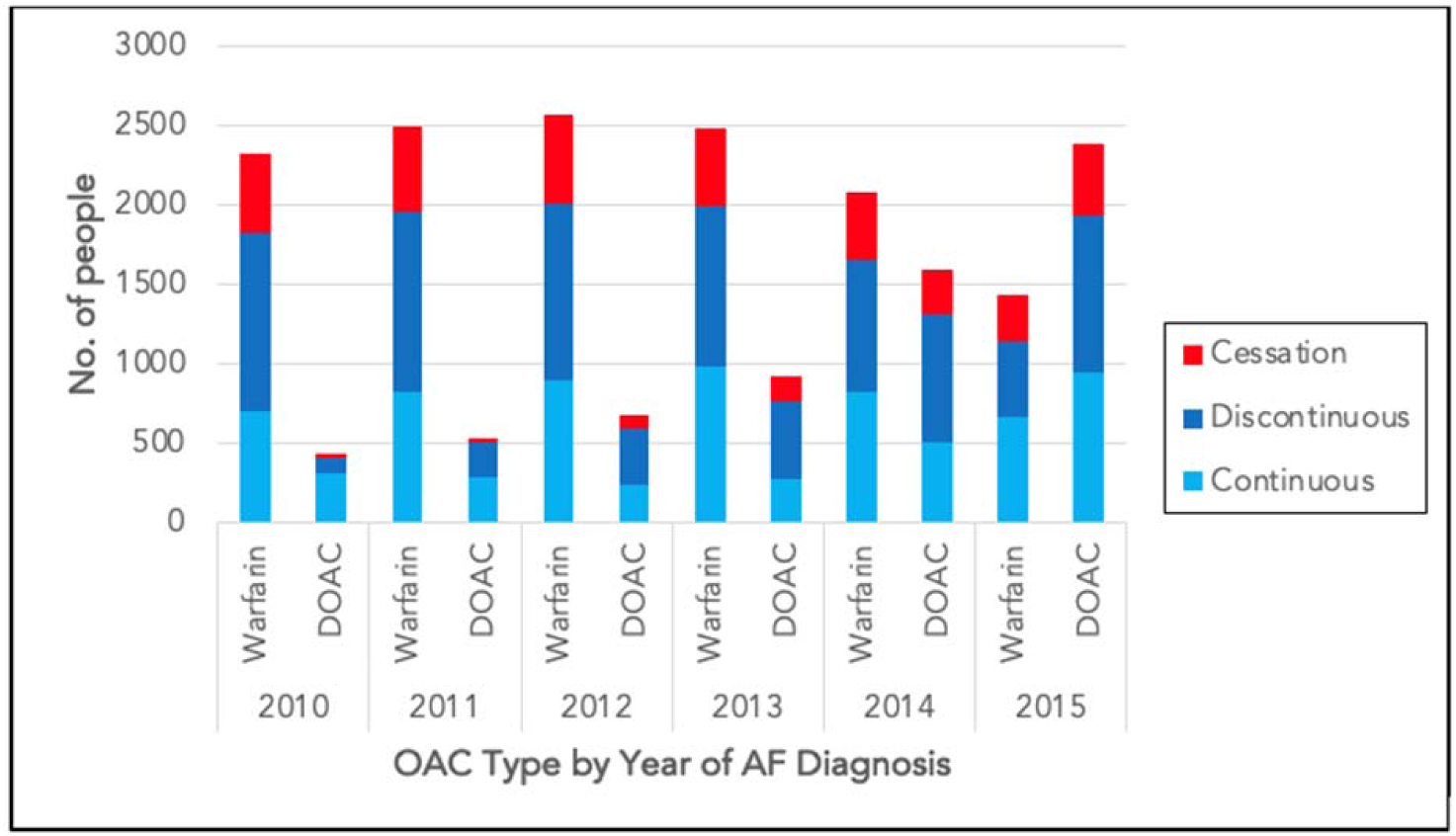
OAC Exposure by OAC Type and Year of AF Diagnosis

### SSE Risk

The percentage diagnosed with stroke or systemic embolism during the five-years after AF diagnosis was highest in the discontinuous OAC therapy cohort (16.54%) and lowest in those that received continuous anti-coagulation (5.99%).

People receiving discontinuous OAC therapy had an increased risk of stroke during the five year follow-up period from an initial AF diagnosis, compared to those that never started oral anti-coagulation (SHR 3.03; 2.81-3.26)(Figure 4A)and those with continuous anti-coagulant prescriptions (SHR 2.65; 2.39-2.94)(Figure 4B). Cessation was associated with greater stoke risk than individuals that never started anti-coagulation (SHR 1.22; 1.22-1.38) (Figure 4B), but lower stroke risk than people in the discontinuous OAC therapy cohort (SHR 0.39;.34-0.44)(Figure 4C). There was no significant difference in stroke risk between people that permanently discontinued anti-coagulation and those with continuous OAC therapy. Prior stroke, age greater than 75, co-morbidities, and elevated frailty risk score were significantly associated with a higher risk of stroke. Compared with a reference score of 2, all CHA_2_DS_2_-VASC scores were associated with a greater risk of SSE. Compared to the most deprived SIMD quintile, the second and third quintiles had similar stroke risk, whilst the fourth and fifth quintiles were associated with a reduced risk of stroke, which was statistically significant. Mortality risk in this population is reported in Supplementary Materials 7.

**Figure 4.**
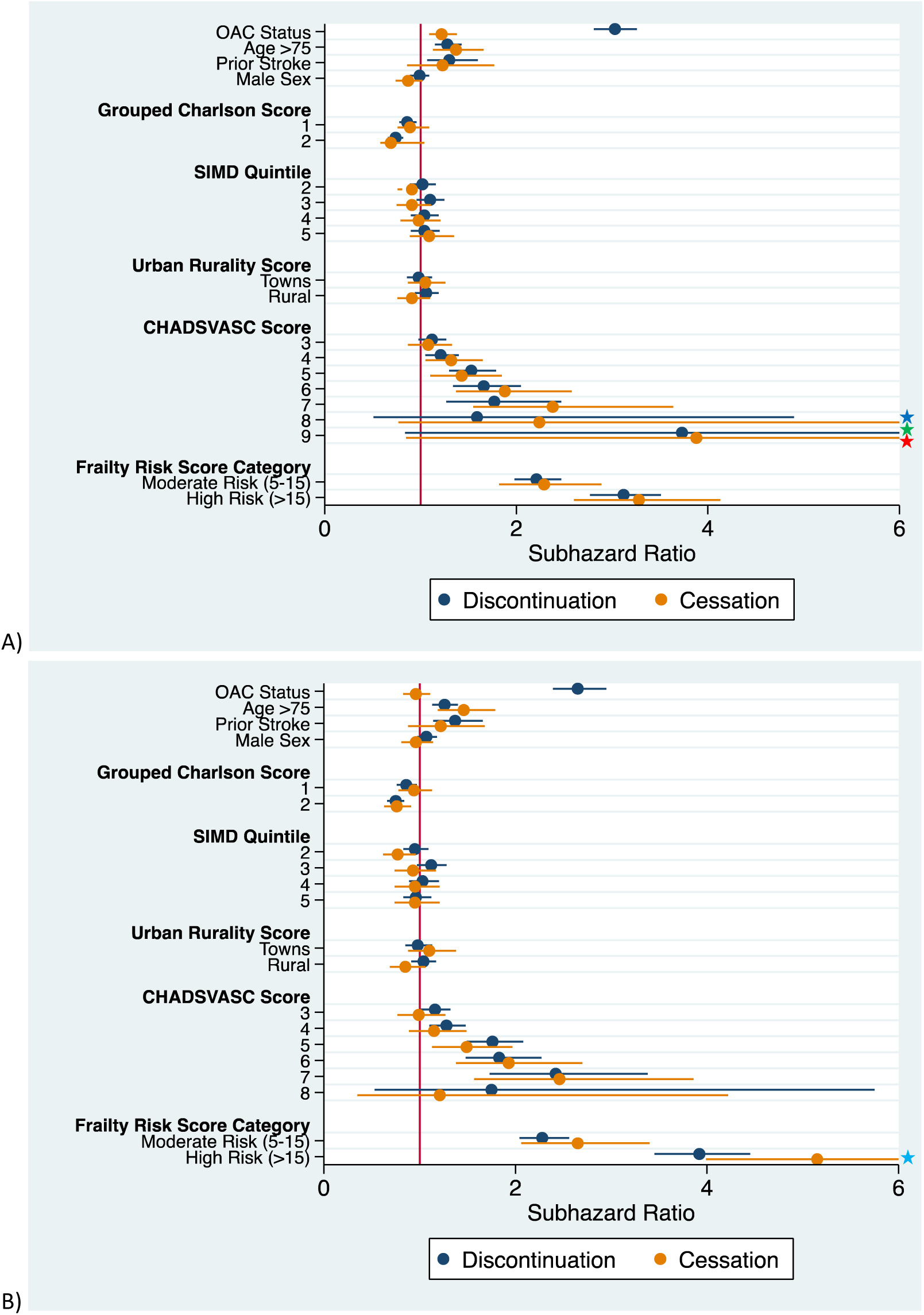
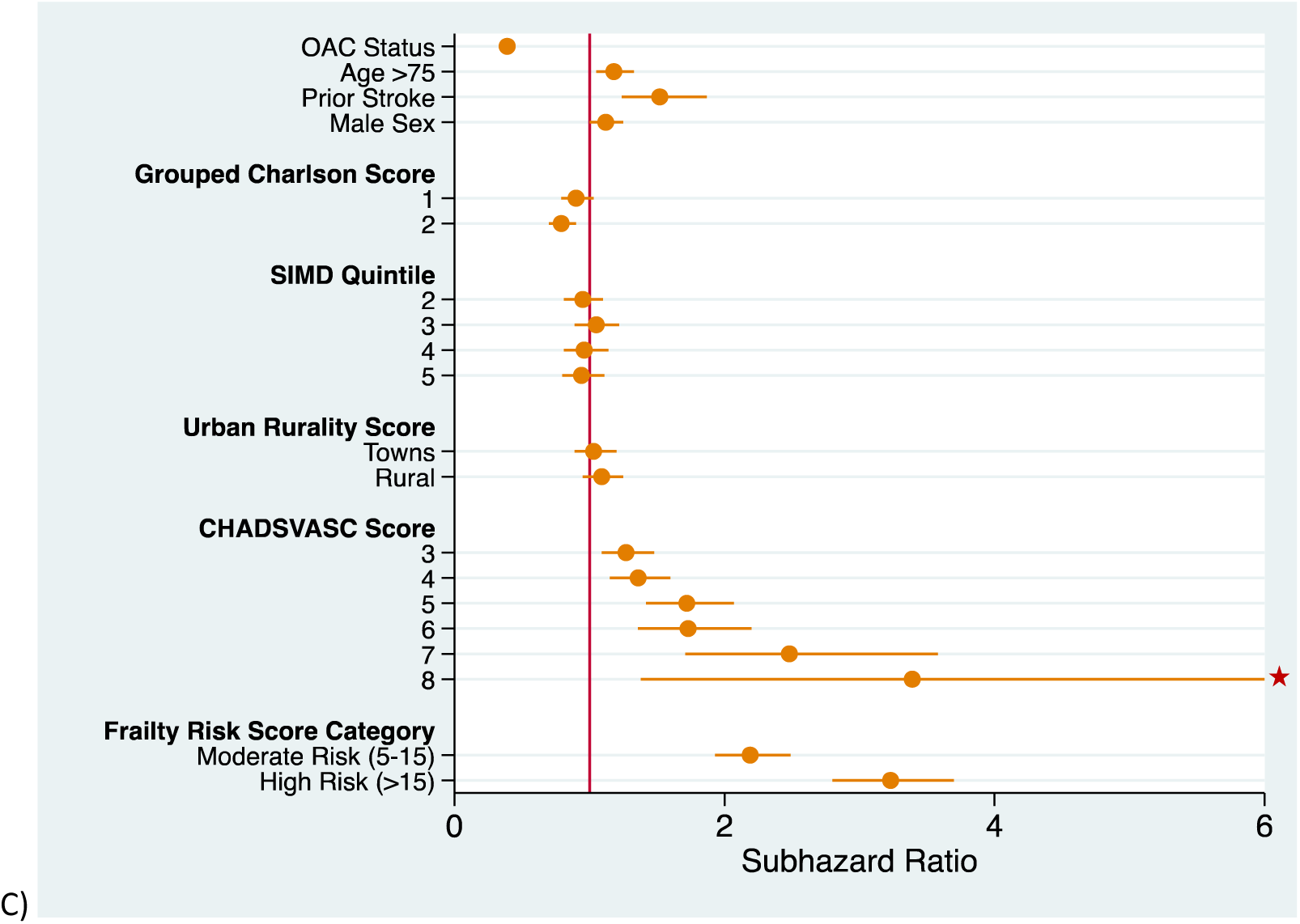
SSE Risk A) Never Started vs Discontinuous and Cessation B) Continuous vs Discontinuous and Cessation C) Discontinuous* vs Cessation *reference Truncated upper confidence intervals: 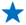16.49 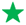6.48 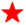17.75 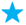6.66 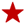8.43

## Major Bleed Analyses

### Anti-coagulation Prescriptions

Warfarin was the most common anti-coagulant in our cohort, prescribed to 71.6% of people. Apixaban was the mostly commonly prescribed DOAC (15%), followed by rivaroxaban (11.5%); dabigatran (1.3%) and edoxaban (0.53%) were utilised more rarely.

### SSE Risk

During five years of follow-up from index AF diagnosis, the highest percentage of stroke and systemic embolism events in individuals with a prior major bleed event, occurred in the discontinuous OAC therapy (33.63%) and cessation (33.33%) cohorts; the continuous OAC therapy population had lowest proportion of stroke events (14.76%).

Discontinuation of OAC therapy was associated with an increased risk of stroke in people that had a prior bleeding event compared to those with continuous OAC therapy (SHR 2.04;.52-2.74)(Figure 4A). Sex, age older than 75, SIMD quintile, urban-rurality classification, time off anti-coagulation and frailty score were not significant mediators of stroke risk in this cohort. Cessation was not associated with a statistically significant difference in risk of stroke compared to those with discontinuous or continuous OAC therapy(Figure 5A/B).

**Figure 5.**
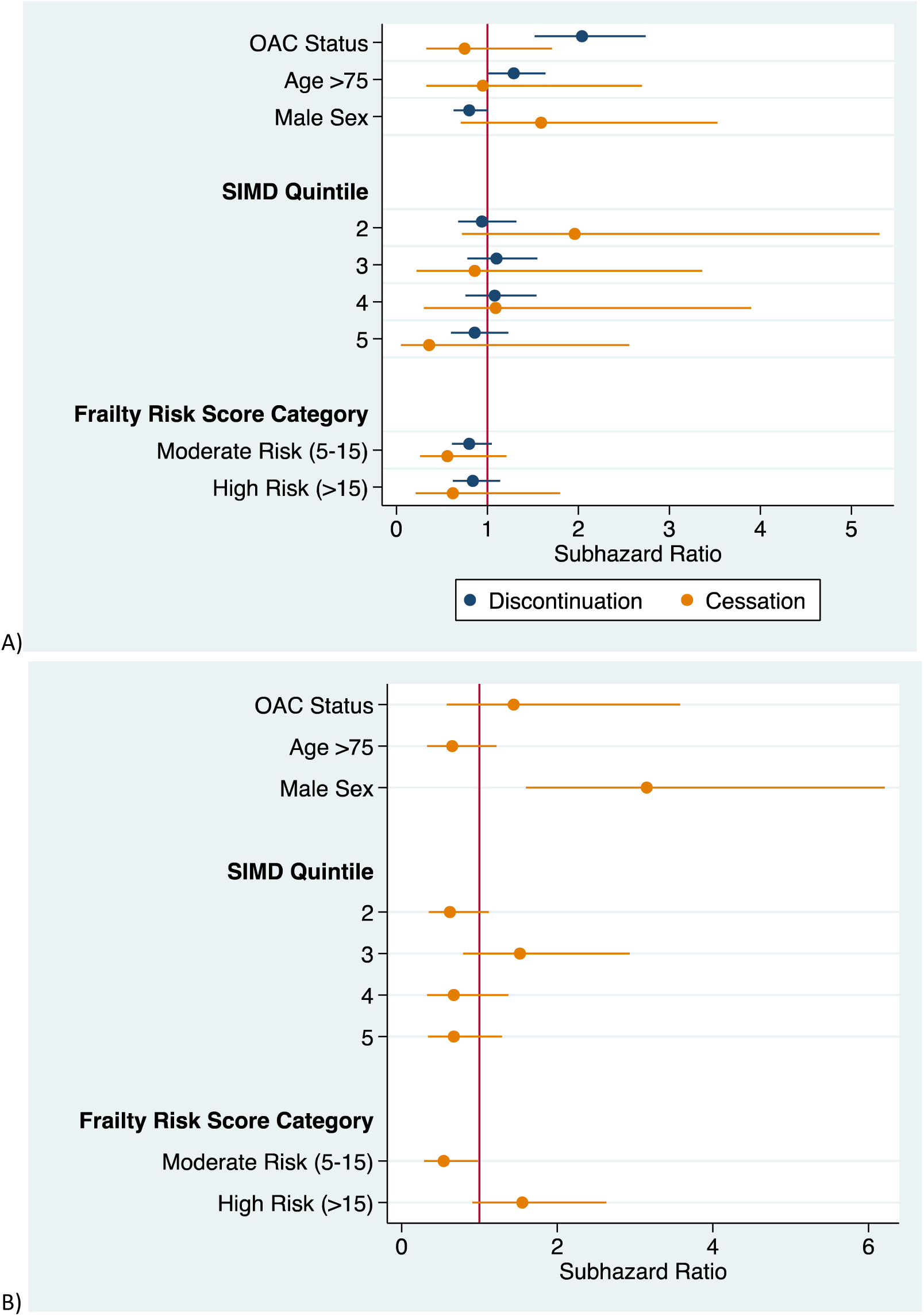
SSE risk following major bleeding event A) Continuous vs Cessation and Discontinuous B) Discontinuous* vs Cessation *reference. Analyses were also adjusted for time off anti-coagulation.

### Bleeding Risk

Recurrent bleeding was most frequent in individuals that discontinued anti-coagulation, either temporarily (27.68%) or permanently (20.5%) or were never prescribed it (22.98%).

The discontinuous OAC therapy cohort had a greater risk of a bleeding event compared to those that persisted with anti-coagulation (SHR 1.71; 95% CI: 1.25-2.33)(Figure 6A). The highest category of frailty risk was associated with an increased risk of recurrent bleeding. Mortality risk in this population is reported in Supplementary Materials 8.

**Figure 6.**
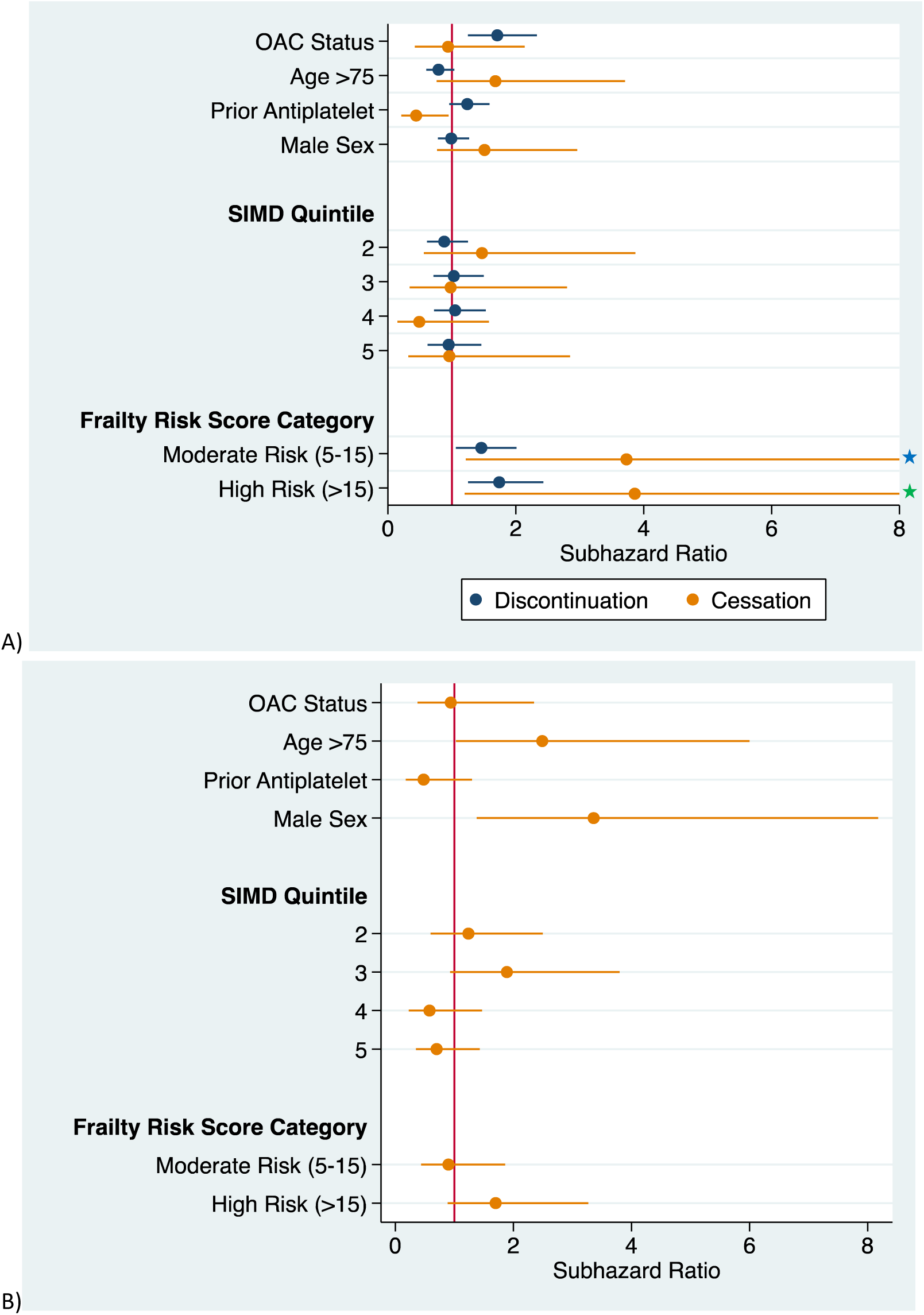
Recurrent bleeding risk A) Continuous vs Cessation and Discontinuous B) Discontinuous* vs Cessation *reference. Truncated upper confidence intervals: 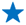11.41 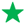12.49 Analyses were also adjusted for time off anti-coagulation.

## Discussion

This retrospective cohort study utilised national linked individual patient data to compare the risks of SSE and bleeding in AF according to anti-coagulant exposure, from which there are multiple key observations. Firstly, despite the high thromboembolic risk in our population, the rate of anti-coagulation was low, and the rate of discontinuation was high, although this improved over the duration of our study. There were considerable inequalities in OAC prescribing; lower socio-economic, elevated frailty score and age ≥75 were associated with anti-coagulation not being commenced and non-continuous OAC prescribing. Furthermore, the SSE risk associated with discontinuation of anti-coagulation appears to be at least equivalent to, if not greater, than never commencing anti-coagulation. Finally, our analyses also indicate that discontinuation or cessation of OACs is not protective with regards to the risk of recurrent bleeding.

### OAC Prescribing

Although our population had an elevated SSE risk, a considerable proportion, 55.4% never commenced warfarin or a DOAC, which is consistent with prior studies highlighting suboptimal anti-coagulation amongst the AF population. Indeed, a recent study by Lee et al. reported that 49.7% of those diagnosed with AF admitted to hospital in Scotland between 2010 and 2019 were not anti-coagulated, although this had improved from 63.8% in 2010 to 35.5% in 2019. (20) They found that women were less likely to be anti-coagulated, although this disparity was mitigated by the increasing prevalence of anti-coagulation with DOACs. Although our data did not include information on the rationale for decision-making on anti-coagulation, De Breucker et al previously reported that the absence of anti-coagulation in older adults is likely due to functional or cognitive impairment, falls risk, malnutrition, or depression. (21)

Whilst sustained endeavours to ensure that anti-coagulation is appropriately initiated at diagnosis are clearly integral, it is also concerning that despite AF being a lifelong condition, requiring ongoing thromboprophylaxis, only 37.51% of our population received continuous OAC therapy. We observed a discontinuation rate of 62.49% with comparable discontinuation between DOACs and warfarin, which is consistent with prior studies. Indeed, a retrospective study by Baker et al. of discontinuation (defined as a temporal gap in prescriptions of greater than thirty days) in 41864 individuals with AF in the USA prescribed DOACs, reported discontinuation rates of 60.3%, 52.8% and 62.9% for rivaroxaban, apixaban and dabigatran respectively.(22) Furthermore, in a retrospective cohort study in the US of 12129 adults with AF, 47% discontinued oral anti-coagulation, which occurred within an average of 120 days; first discontinuation often occurs early after the initial prescription.(23)

Some studies considered adherence and persistence, rather than, or in addition to, discontinuation explicitly, in their analyses. Indeed, Dhamane et al. conducted a retrospective analysis of non-persistence, defined as treatment switching, or a refill-gap of 60 days or greater, in over one million people with AF prescribed anti-coagulants in the US.(24) At one year, the cumulative incidence of non-persistence was 51.3%, 58.9%, 51.3% and 52.2%, for warfarin, apixaban, rivaroxaban and dabigatran, respectively. Few studies have evaluated cessation of anti-coagulation; a prospective cohort study in Italy assessing cessation of DOACs in 1305 adults with AF reported that 15.4% were no longer anti-coagulated after one year of follow-up; greater than 60% discontinued within the initial 6 months. (25)

Our analyses also suggest considerable inequalities in OAC prescribing, which are essential to address, given that non-initiation and discontinuity of anti-coagulation are associated with poorer clinical outcomes, including a greater SSE risk . For example, those living in the most deprived areas were most likely to have never commenced anti-coagulation, whilst those living in the most affluent areas were most likely to have been prescribed anti-coagulation. A previous study of stroke survivors with AF in Scotland found that the most deprived SIMD quintiles were associated with an absence of anti-coagulation. (26) Furthermore, those in the least deprived quintile were most likely to have continuous anti-coagulation, which is consistent with international studies(27, 28). However, in a recent study of trends in OAC prescribing in England, a higher socio-economic status appeared to be associated with non-adherence.(29)

### Stroke Risk Following OAC Discontinuation

Our analyses indicate that OAC discontinuation is associated with a significantly increased SSE risk in adults with AF, compared to those that are continuously anti-coagulated. Indeed, Toorop et al. evaluated OAC persistence in a Dutch AF cohort, defined as a gap of less than one hundred days between the final OAC prescription and the study end date, and found that non-persistence was associated with a 58% increase in the risk of ischaemic stroke compared to those with continuous OAC exposure.(30) Similarly, Rodriguez et al. conducted a nested case-control analysis of electronic health records in the UK and Denmark, matching incident cases of ischaemic stroke to controls by age and sex, and found that those that discontinue OAC have a risk of stroke two to three times greater than those with continuous treatment.(31)

In our population, temporary discontinuation of anti-coagulation was also associated with a greater thromboembolic risk than in people that never initiated OAC therapy, and in those that permanently discontinued. Discontinuation is ostensibly associated with a period of significant clinical risk during which individuals require active, dynamic monitoring. Indeed, rebound hypercoagulability has been postulated to occur transiently in the aftermath of discontinuation, associated with raised blood markers of thrombin production.(32) A cohort study in South Korea reported that discontinuation of DOACs was associated with greater stroke severity at initial presentation, per the National Institute of Health Stroke Scale (NIHSS), than those that discontinued warfarin and those never commenced on anti-coagulation which they postulated was due to this hypercoagulable state.(33)

### Outcomes Following OAC Discontinuation in the Context of a Major Bleeding Event

This study found that, in the context of a major bleeding event, individuals that discontinued anti-coagulation had an increased risk of SSE and of a further bleeding event compared to those continuously anti-coagulated. Furthermore, our analyses indicated no significant difference in the risk of a subsequent bleed between those that permanently discontinued compared to those that continued anti-coagulation, in those who had experienced a major bleeding event. Ewen et al. previously found no difference in rates of bleeding events between people with continuous and discontinuous anti-coagulation, although this was in a primary care setting with a shorter follow-up duration of twelve months. (34) Several prior cohort studies, and a meta-analysis, have reported that recommencing anti-coagulation following a bleed is associated with a lower risk of ischaemic stroke and all-cause mortality to those for whom anti-coagulation remained withheld, with no statistically significant difference in the risk of bleeding. (5, 35–37) However, these have typically focused on single type of bleeding event, for example, gastrointestinal bleeding, or only considered these outcomes in the short-term. Consensus on the optimum time to reinitiate anti-coagulation following a major bleeding event is currently lacking; the American Heart Association recommends re-initiation of anti-coagulation within 7-10 days following an intracranial haemorrhage, whilst others have proposed 10 weeks.(38) Acutely, the risk of further bleeding is likely greater than the risk of stroke or systemic embolism, whilst in the longer-term risk of thromboembolism may exceed that of a recurrent bleed.

### Strengths and Limitations

A key strength of our study is the use of national linked data to understand, over a relatively long follow-up period, prescribing patterns in people with AF, and effects of discontinuation of anti-coagulation, which could not feasibly be investigated within the context of an RCT. Our results are generalisable to similar AF populations.

However, our study has several possible limitations. Firstly, since SMR01 and SSCA only include diagnoses relevant to hospital admissions, AF diagnoses in primary care were not captured in our data.(39) Furthermore, our secondary care AF population is likely at greater risk of the outcomes measured than people managed exclusively in primary care.

Second, clinical miscoding is a possible risk of utilising data derived from administrative healthcare records. Furthermore, PIS only captures primary care prescribing data, such that secondary care prescriptions of anti-coagulants may thus create a temporal gap within the PIS data. A refill gap of 60 days was thus selected to afford sufficient time for community prescribing to have resumed following a secondary care admission, to reduce the potential for erroneous assignment of people to the discontinuous cohort following hospital discharge, since the reason for discontinuation is not recorded in PIS.

Finally, given the heterogeneous patterns of adherence for anti-coagulation, our definition for discontinuation may not have adequately captured the risks of our outcome measures for all individuals in this cohort. Indeed, individuals within this cohort may have had significantly different time periods in which they were not anti-coagulated.

## Future Research

Harmonisation of definitions and methodologies in the evaluation of discontinuation of anti-coagulation would be highly valuable in aiding comparisons since there is significant heterogeneity in the existing literature.

Given the under-utilisation of thromboprophylaxis, and high discontinuation rates observed in our analyses and in the wider literature, further research is critical to elucidate the rationales underlying decision-making more clearly, by clinicians and patients, around anti- coagulant prescribing. Indeed, this will allow the development of targeted interventions aimed at promoting appropriate OAC initiation on diagnosis of AF and mitigating OAC discontinuation, such that stroke prevention may be optimised; patient-centred approaches which promote shared decision-making between clinicians and patients are valuable in supporting adherence.(40) This is particularly important since there is increasing evidence that the lack of monitoring for DOACs, whilst convenient, may be deleterious to persistence; monitoring appointments afford healthcare professionals the opportunity for continued patient education to reinforce the rationale for anti-coagulation.(41)

Further research is necessary to establish an optimal approach to management of anti-coagulants in adults with AF in the context of major bleeding events, given the lack of consensus amongst clinicians, and consequent variation in real-world clinical practice. This would support the identification of individuals who may safely recommence anti-coagulation at the appropriate timepoint following a bleed, as well as those in whom future anti-coagulation is potentially contra-indicated, optimising clinical outcomes.

## Conclusion

Considerable inequalities in OAC prescribing exist for people with AF in Scotland, which is of particular significance since non-initiation and discontinuity of anti-coagulation are associated with poorer clinical outcomes, including increased SSE risk. Given the considerable clinical risks associated with OAC discontinuation reported in this study and in the wider literature, active monitoring is necessary. Decision-making on OAC prescribing is complex, but to optimise clinical outcomes, it must be patient-centred, and account for thrombotic and bleeding risks, particularly where these differ from the baseline risks that informed the initial prescription.

## Acknowledgements

We acknowledge the invaluable support of the staff at PHS.

